# Effects of testing and vaccination levels on the dynamics of the COVID-19 pandemic and the prospects for its termination

**DOI:** 10.1101/2021.09.20.21263823

**Authors:** Igor Nesteruk, Oleksii Rodionov

## Abstract

A simple statistical analysis of the accumulated and daily numbers of new COVID-19 cases and deaths per capita was performed with the use of recent datasets for European and some other countries and regions. It was shown that vaccination can significantly reduce the likelihood of deaths. However, existing vaccines do not prevent new infections, and vaccinated individuals can spread the infection as intensely as unvaccinated ones. Therefore, it is too early to lift quarantine restrictions in Europe and most other countries. The constant appearance of new cases due to re-infection increases the likelihood of new coronavirus strains, including very dangerous. As existing vaccines are not able to prevent this, it remains to increase the number of tests per registered case. If the critical value of 520 is exceeded, one can hope to stop the occurrence of new cases.

## Introduction

To investigate the effectiveness of quarantine, testing and vaccination, different relative characteristics (calculated per capita) can be used. In particular, such values are regularly reported by COVID-19 Data Repository by the Center for Systems Science and Engineering (CSSE) at Johns Hopkins University (JHU), [1]. The accumulated numbers of COVID-19 cases per capita (CC) was used in [2] to investigate the influence of demographic factors in European countries. In the end of June 2021 the CC values varied more than 9 times for different European countries but showed no visible dependencies on the volume of population, its density, and the level of urbanization. In this paper we will try to find some statistical correlation between CC values and the accumulated number of tests per capita (TC) and the tests per cases ratio TC/CC. We will try also to find similar relationships for the accumulated numbers of deaths per capita caused by coronavirus (DC) and the mortality rate DC/CC versus TC and TC/CC.

The current dynamics of the pandemic is characterized by daily increases in the number of cases. The daily numbers of new COVID-19 cases per capita (DCC) was used in [3] to find some seasonal trends of the COVID-19 pandemic in the EU and some other countries. This characteristic and the daily numbers of new deaths per capita caused by coronavirus (DDC) are very important in order to investigate the efficiency of vaccinations. In particular, it was shown in [4] that rather high numbers of fully vaccinated people per capita (VC) did not protect the population of Israel against a new pandemic wave in the summer of 2021. In this paper we will try to find a correlation between DCC, DDC and VC values.

## Data, the linear regression and Fisher test

We will use the data sets regarding the relative characteristics (per capita) reported by JHU as of September 1, 2021, [1]. The figures corresponding to the version of the JHU table available on September 12, 2021 are presented in Table 1. We cannot fix the date September 12 for all the data, since many figures appear in the JHU table with the delay. The accumulated characteristics: CC (number of cases per million), DC (number of deaths per million), TC (number of tests per thousand), VC (percentage of fully vaccinated people) are taken without smoothing.

**Table 1.** Accumulated and daily characteristics of the COVID-19 pandemic dynamics in European and some other countries and regions as of September 1, 2021 (figures corresponding to other days in May-September 2021 are specified in notes), [1].

Data, the linear regression and Fisher test
| Country | Total cases<br>per million<br>CC | Total death<br>per million<br>DC | Total tests<br>per thousand<br>TC | People fully<br>vaccinated,<br>%,<br>VC | New cases<br>per million,<br>smoothed<br>DCC | New deaths<br>per million,<br>smooth.<br>DDC |
| --- | --- | --- | --- | --- | --- | --- |
| **European countries** |  |  |  |  |  |  |
| Monaco | 81250 | 835.02 | no data | 57.07<sup>1</sup> | 173.511 | 0 |
| Holy See (Vatican City) | 33251.232 | no data | no data | no data | 0 | 0 |
| Malta | 70442.161 | 857.036 | 2321.107 | 80.27 | 94.116 | 1.111 |
| San Marino | 156748.02 | 2646.281 | no data | 70.49 | 163.817 | 0 |
| Netherlands | 115262.92 | 1069.871 | 736.14<sup>2</sup> | 62.64<sup>2</sup> | 152.897 | 0.516 |
| Belgium | 102086.65 | 2182.021 | 1614.388 | 70.14 | 176.822 | 0.418 |
| United Kingdom | 100531.05 | 1950.911 | 3590.956 | 63.08 | 492.479 | 1.556 |
| Liechtenstein | 85690.385 | 1542.322 | 1624.614 | 53.55 | 115.768 | 0 |
| Luxembourg | 119568.88 | 1307.47 | 5400.155 | 56.18 | 133.222 | 0 |
| Germany | 47435.24 | 1099.66 | 838.014<sup>2</sup> | 60.34 | 116.255 | 0.293 |
| Italy | 75313.524 | 2141.716 | 1397.101 | 61.1 | 104.34 | 0.89 |
| Switzerland | 89824.971 | 1251.449 | 1079.174 | 51.23 | 289.779 | 0.82 |
| Andorra | 194508.36 | 1680.585 | 2709.918<sup>2</sup> | 54.08 | 59.098 | 0 |
| Denmark | 59838.969 | 444.842 | 6874.115 | 72.59 | 147.027 | 0.319 |
| Czech Republic | 156601.03 | 2834.99 | no data | 53.6 | 18.556 | 0.226 |
| Poland | 76435.59 | 1993.756 | 513.568 | 49.8 | 6.675 | 0.11 |
| Portugal | 102232.48 | 1746.374 | 1682.166 | 75.37 | 191.4 | 1.166 |
| Slovakia | 72357.046 | 2297.863 | 7527.671 | 39.81 | 21.897 | 0.026 |
| Albania | 51295.644 | 870.539 | 256.537<sup>3</sup> | 22.58 | 298.55 | 0.895 |
| Austria | 76318.424 | 1191.52 | 8492.748 | 57.82 | 157.563 | 0.079 |
| France | 101653.62 | 1702.542 | no data | 60.19 | 233.095 | 1.672 |
| Hungary | 84338.524 | 3120.043 | 636.628 | 57.2 | 17.705 | 0.059 |
| Turkey | 75400.291 | 670.251 | 901.9 | 43.94 | 232.817 | 3.004 |
| Slovenia | 128907.03 | 2140.737 | 699.719 | 43.58 | 227.681 | 0.481 |
| Moldova | 66626.077 | 1591.938 | 393.622<sup>1</sup> | 17.29 | 81.226 | 0.923 |
| Spain | 104008.15 | 1807.073 | 1186.821<sup>4</sup> | 72 | 142.652 | 2.39 |

|  | CC | DC | TC | VC | DCC | DDC |
| --- | --- | --- | --- | --- | --- | --- |
| Serbia | 112611.98 | 1073.833 | 741.032 | 41.31 | 369.204 | 1.554 |
| Romania | 57518.879 | 1808.418 | 477.155<sup>1</sup> | 26.89 | 54.05 | 1.031 |
| North Macedonia | 85179.009 | 2863.644 | 553.002<sup>5</sup> | 25.88 | 399.42 | 14.336 |
| Greece | 56971.017 | 1318.034 | 1506.965 | 55.43 | 285.9 | 2.893 |
| Bosnia and Herzegovina | 65807.17 | 3007.545 | 350.723 | 13.06<sup>9</sup> | 161.967 | 2.495 |
| Croatia | 91826.187 | 2042.798 | 623.275<sup>6</sup> | 39.68 | 133.629 | 0.98 |
| Ireland | 71090.272 | 1025.908 | 1348.36 | 68.09 | 341.367 | 0.573 |
| Ukraine | 54916.644 | 1312.863 | 276.267 | 8.99 | 49.815 | 1.479 |
| Bulgaria | 66334.622 | 2747.709 | 620.083<sup>1</sup> | 17.12 | 223.483 | 6.981 |
| Belarus | 51174.183 | 401.467 | 827.889 | 14.15<sup>2</sup> | 157.02 | 1.195 |
| Montenegro | 184628.32 | 2757.738 | no data | 29.45 | 959.886 | 8.416 |
| Lithuania | 111355.9 | 1698.6 | 1723.318 | 56.23 | 220.882 | 3.399 |
| Latvia | 76652.951 | 1381.409 | 1788.741<sup>7</sup> | 40.96 | 120.136 | 0.689 |
| Estonia | 107428.53 | 975.711 | 1335.17 | 41.27 | 261.526 | 0.862 |
| Sweden | 111013.72 | 1446.04 | no data | 56.21<sup>2</sup> | 102.487 | 0.253 |
| Norway | 29605.742 | 150.394 | 1318.59 | 58.07 | 253.402 | 0.209 |
| Finland | 23044.824 | 185.64 | 1201.377 | 51.1 | 108.809 | 0.309 |
| Iceland | 31616.962 | 96.109 | 1715.983 | 76.82 | 214.269 | 1.248 |
| **Other countries and regions** |  |  |  |  |  |  |
| United States | 118336.96 | 1929.465 | 1604.422 | 51.91 | 503.381 | 4.186 |
| North America | 79327.442 | 1643.263 | no data | 42.43 | 338.245 | 3.973 |
| Argentina | 113822.04 | 2455.936 | 293.9 | 32.42 | 112.357 | 3.255 |
| Brazil | 97218.938 | 2715.737 | 148.213<sup>8</sup> | 29.68 | 105.93 | 3.007 |
| South America | 85023.252 | 2605.171 | no data | 31.06 | 76.457 | 2.336 |
| South Africa | 46420.892 | 1373.972 | 275.411 | 10.23 | 154.656 | 4.823 |
| Africa | 5694.404 | 143.37 | no data | 2.86 | 20.617 | 0.535 |
| India | 23580.97 | 315.434 | 375.471 | 10.9 | 30.696 | 0.324 |
| Israel | 123209.17 | 806.164 | 2675.281 | 62.52 | 1058.958 | 2.893 |
| Japan | 11991.372 | 128.131 | 166.789 | 46.85 | 161.421 | 0.434 |
| South Korea | 4978.074 | 44.888 | 241.129 | 31.74 | 33.647 | 0.128 |
| Qatar | 79477.254 | 205.424 | 856.443 | 73.44 | 65.859 | 0.049 |
| Asia | 15035.556 | 222.955 | no data | 28.91 | 54.181 | 0.868 |
| Australia | 2193.25 | 39.514 | 1233.936 | 28.37 | 48.306 | 0.166 |
| World | 27737.424 | 575.503 | no data | 27.3 | 81.926 | 1.232 |

Figures corresponding different days in 2021: <sup>1</sup> 09-02; <sup>2</sup> 08-29; <sup>3</sup> 06-13; <sup>4</sup> 08-26; <sup>5</sup> 08-30; <sup>6</sup> 08-31; <sup>7</sup> 08-27; <sup>8</sup> 05-24; <sup>9</sup> 09-07.

Since daily characteristics DCC (new cases per million) and DDC (new deaths per million) are very random and demonstrate some weekly periodicity, we will use smoothed (averaged) values calculated and displayed by JHU with the use of figures registered during the previous 7 days. This smoothing procedure differs from one proposed in [5-7], where the values registered in the nearest 7 days were used.

We will use the linear regression to calculate the regression coefficients *r* and the coefficients *a* and *b* of corresponding best fitting straight lines, [8]:

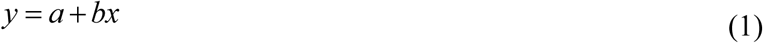

where *x* are TC, TC/CC, and VC values and *y* are CC, DC, DC/CC, DCC, DDC, and DDC/DCC values.

We will use also the F-test for the null hypothesis that says that the proposed linear relationship (1) fits the data sets. The experimental values of the Fisher function can be calculated with the use of the formula:

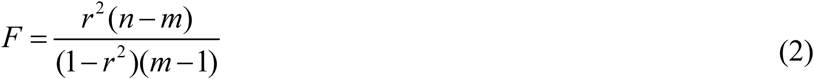

where *n* is the number of observations (number of countries and regions taken for statistical analysis); *m*=2 is the number of parameters in the regression equation, [8]. The corresponding experimental values *F* have to be compared with the critical values *F*_*C*_ (*k*_1_, *k*_2_) of the Fisher function at a desired significance or confidence level (*k*_1_ = *m* −1, *k*_2_ = *n* − *m*, see, e.g., [9]). If *F*_*C*_ (*k*_1_, *k*_2_) / *F* < 1, the null hypothesis is not supported by the results of observations. The highest values of *F*_*C*_ (*k*_1_, *k*_2_) / *F* correspond to the most reliable hypotheses (see, e.g., [10]).

## Results

The results of the linear regression application are presented in Table 2 and Figs. 1-3. We have used the data sets corresponding to the European countries and complete datasets with the information about some other countries and regions. Thus, we have two different numbers of observations *n* for every application of the linear relationship (1). Due to the lack of some data, the numbers *n* are different for different relationships (e.g., versus TC and versus VC). Corresponding values of the regression coefficients *r*, coefficients *a* and *b*; values of the Fisher function *F, F*_*C*_ (*k*_1_, *k*_2_) and *F* / *F*_*C*_ (*k*_1_, *k*_2_) are shown in Table 2. The best fitting lines (1), calculated with the use of corresponding values *a* and *b* are shown in Figs. 1-3 by solid lines for European datasets and by dashed lines for complete datasets. The values CC, DC, CC/DC, DCC, DDC, and DDC/DCC are represented by “crosses” for European countries and by “circles” for other countries and regions.

**Table 2.** Optimal values of parameters in eq. (1), correlation coefficients and the results of Fisher test applications.

| Number of Figure, relationship | Number of observations<br>$n$ | Correlation coefficient<br>$r$ | Optimal values of parameters<br>$a$<br>in eq. (1) | Optimal values of parameters<br>$b$<br>in eq. (1) | Experimental value of the Fisher function $F$,<br>eq. (2),<br>$m=2$ | Critical value of Fisher Function<br>$F_c(1, n-2)$<br>for the confidence level 0.1, [9] | $F_c/F$ |
| --- | --- | --- | --- | --- | --- | --- | --- |
| 1, CC versus TC | 37 | 0.0952 | 7.8115e+04 | 1.5147 | 0.3202 | 2.86 | 0.1120 |
|  | 47 | 0.1728 | 7.1474e+04 | 3.3914 | 1.3848 | 2.84 | 0.4876 |
| 1, DC versus TC | 37 | -0.1176 | 1.5699e+03 | -0.0454 | 0.4906 | 2.86 | 0.1715 |
|  | 47 | -0.0478 | 1.4190e+03 | -0.0218 | 0.1031 | 2.84 | 0.0363 |
| 1, DC/CC versus TC | 37 | -0.1793 | 0.0205 | -9.1515e-07 | 1.1620 | 2.86 | 0.4063 |
|  | 47 | -0.1565 | 0.0194 | -8.3788e-07 | 1.1293 | 2.84 | 0.3977 |
| 2, CC versus TC/CC | 37 | -0.2471 | 8.7751e+04 | -2.7460e+05 | 2.2752 | 2.86 | 0.7955 |
|  | 47 | -0.3580 | 8.2349e+04 | -1.5841e+05 | 6.6143 | 2.84 | 2.3290 |
| 2, DC versus TC/CC | 37 | -0.3191 | 1.7042e+03 | -8.6173e+03 | 3.9668 | 2.86 | 1.3870 |
|  | 47 | -0.3161 | 1.4969e+03 | -3.2532e+03 | 4.9951 | 2.84 | 1.7588 |
| 2, DC/CC versus TC/CC | 37 | -0.2460 | 0.0210 | -0.0877 | 2.2544 | 2.86 | 0.7883 |
|  | 47 | -0.0790 | 0.0184 | -0.0095 | 0.2826 | 2.84 | 0.0995 |
| 3, DCC versus VC | 43 | -0.1004 | 234.6744 | -0.8581 | 0.4171 | 2.85 | 0.1464 |
|  | 58 | 0.1245 | 136.6035 | 1.2095 | 0.8821 | 2.8 | 0.3150 |
| 3, DDC versus VC | 43 | -0.3730 | 4.1071 | -0.0521 | 6.6256 | 2.85 | 2.3248 |
|  | 58 | -0.2972 | 3.2549 | -0.0359 | 5.4255 | 2.8 | 1.9377 |
| 3, DDC/DCC versus VC | 43 | -0.5087 | 0.0194 | -2.3335e-04 | 14.3156 | 2.85 | 5.0230 |
|  | 58 | -0.5855 | 0.0227 | -2.8822e-04 | 29.2106 | 2.8 | 10.4324 |

**Fig. 1.**
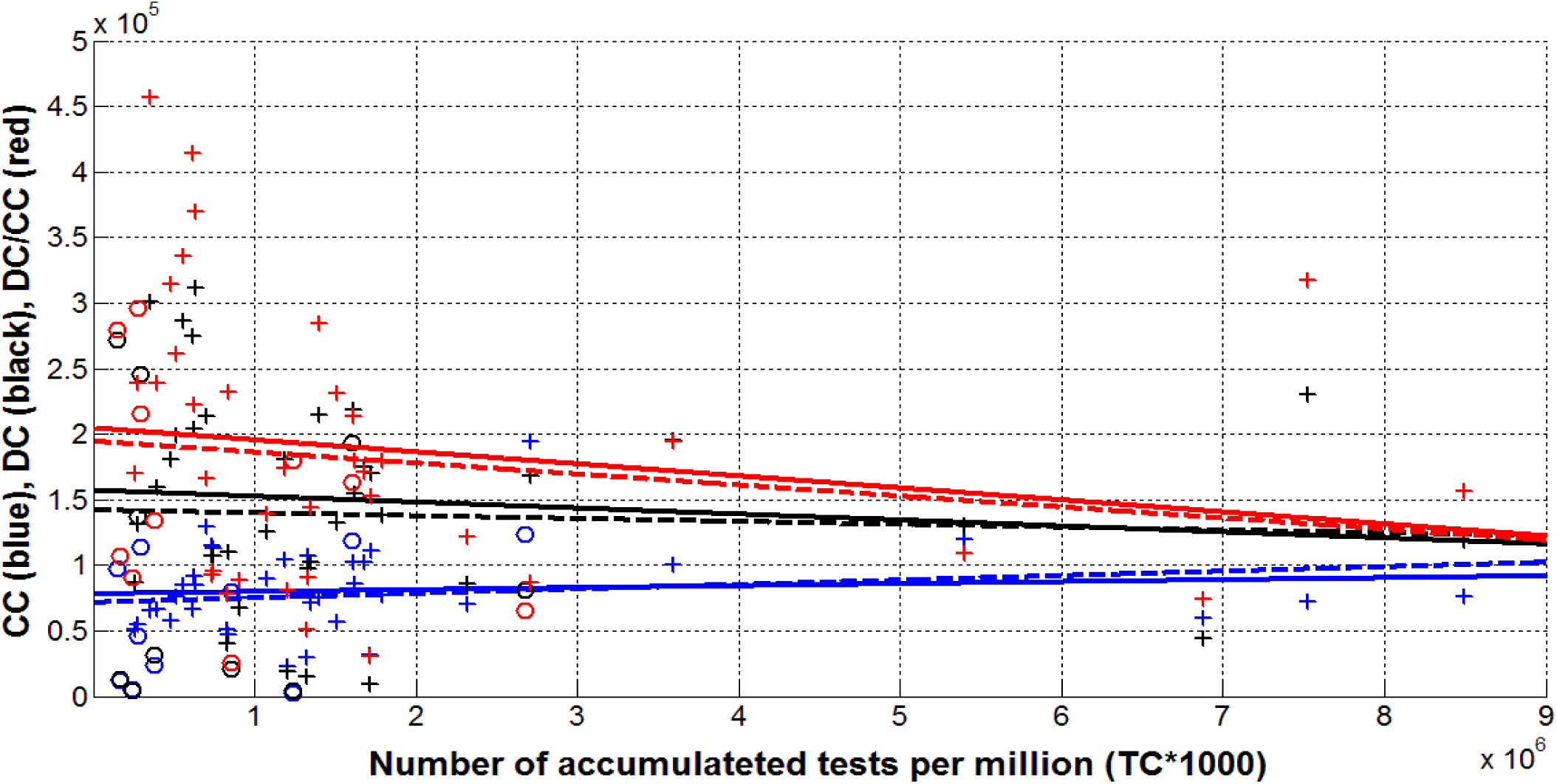
Characteristics of the COVID-19 pandemic versus number of tests per million (TC*1000) accumulated up to September 1, 2021. Cases per million (CC, blue), deaths per 100 million (DC*100, black), mortality rate (DC*10^7^/CC, red). Best fitting lines (1) are solid for European datasets and dashed for complete datasets. “Crosses” represent the European datasets, “circles” – figures for other countries and regions.

**Fig. 2.**
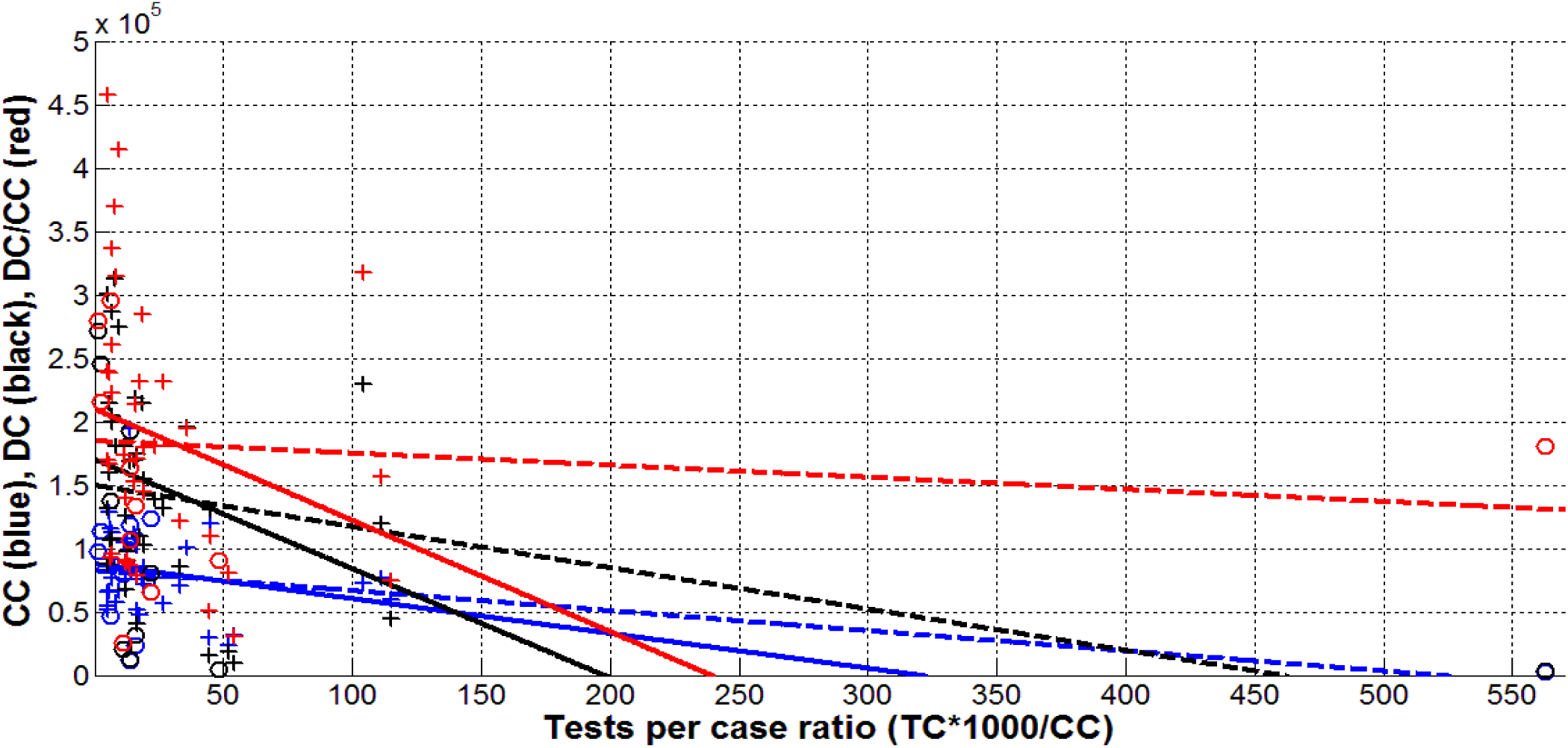
Characteristics of the COVID-19 pandemic versus tests per case ratio (TC*1000/CC) accumulated up to September 1, 2021. Cases per million (CC, blue), deaths per 100 million (DC*100, black), mortality rate (DC*10^7^/CC, red). Best fitting lines (1) are solid for European datasets and dashed for complete datasets. “Crosses” represent the European datasets, “circles” – figures for other countries and regions.

**Fig. 3.**
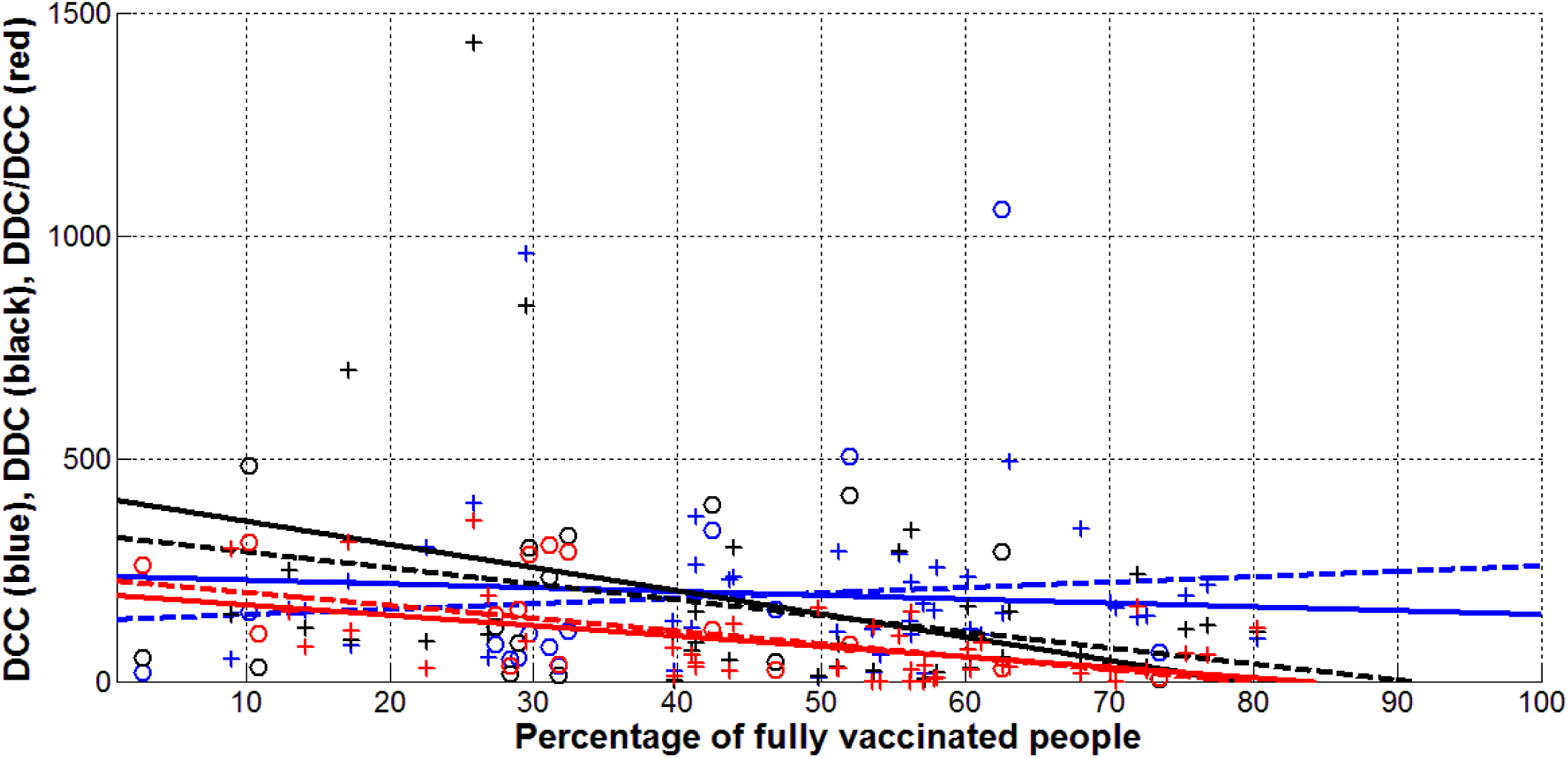
Smoothed daily characteristics of the COVID-19 pandemic versus percentage of fully vaccinated people (VC) as of September 1, 2021. New cases per million (DCC, blue), new deaths per 100 million (DDC*100, black), daily mortality rate (DDC*10^4^/DCC, red). Best fitting lines (1) are solid for European datasets and dashed for complete datasets. “Crosses” represent the European datasets, “circles” – figures for other countries and regions.

Table 2 and Fig. 1 illustrate that there is no visible correlation between all the relative accumulated characteristics (CC, DC and DC/CC) and the accumulated number of tests per capita (TC), since *F* / *F*_*C*_ (*k*_1_, *k*_2_) < 1 for all 6 calculations (see the rows in Table 2 corresponding to Fig.1). Statistical analysis did not confirm the common view that more tests per capita can better identify patients and lead to an increase in CC and DC values. A very weak growth trend with increasing number of tests we see for CC values (see blue lines in Fig. 1). For values DC and DC/CC, black and red lines illustrate opposite trends. This probably triggers the fact that more tests can detect and isolate patients more quickly, slowing the spread of infection and the number of deaths. The competition of these two tendencies results in an almost imperceptible correlation. It looks that it is impossible to stop the pandemic by increasing the number of tests per capita.

In comparison, the increase of the relative characteristic - number of tests per case ratio (TC*1000/CC) – always diminishes the CC, DC and DC/CC values (see lines in Fig. 2). Moreover, visible correlations with TC/CC were revealed for CC and DC values (*F* / *F*_*C*_ (*k*_1_, *k*_2_) < 1 only in the relationship CC versus TC/CC for European countries). This fact allows us to conclude that the increase in the tests per case ratio could stop the pandemic. To calculate the corresponding TC/CC values lets us put in eq. (1) *y=0* and obtain the corresponding *x*_*0*_ value as follows:

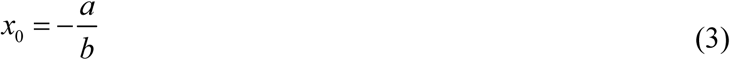

Application of formula (3) for 3 cases with *F* / *F*_*C*_ (*k*_1_, *k*_2_) > 1, yields the result that new COVID-19 cases in the world could stop when TC*1000/CC > 520 (see the blue dashed line in Fig. 2); deaths in the world could disappear at test per case ratio greater than 460 (see the black dashed line); the critical value for the deaths in Europe is TC*1000/CC = 198 (see the black solid line).

Only one country – Australia – exceeded these estimations for the critical values of the tests per case ratio (see last right “circles” in Fig. 2). The corresponding CC and DC values in this country are very low (see blue and black “circles”). Nevertheless, new cases and deaths occur in Australia. This situation can be explained by a decrease in the number of tests. As of September 1, the daily number of tests per 1,000 population was 10,069 [1]. Taking the corresponding DCC value 48.306 from Table 1, we obtain 208.4 as a recent value of the tests per case ratio.

In contrast to the values of DC, the ratio DC/CC does not show any visible correlation with the tests to case ratio TC*1000/CC especially for the complete datasets (in Table 2, the corresponding value of the regression coefficient *r* is -0.0790). This statistical conclusion may seem strange at first glance, but the ratio DC/CC shows how many sick people die. That is, it is a characteristic of the ability of patients to resist the captured strain of the coronavirus and the level of medical care. Therefore, it should not depend on the number of tests performed to identify one infected patient.

The effect of vaccination levels VC on the daily number of new cases DCC was much unexpected due to the practical lack of correlation. We can see in Table 1 the values of the correlation coefficients -0.1004 and 0.1245 for the European and complete datasets, respectively and *F* / *F*_*C*_ (*k*_1_, *k*_2_) << 1. The same conclusion can be drawn from the best fitting blue lines shown in Fig. 3. The available statistical data show that new cases will appear even if the entire population of the earth is vaccinated. But vaccination significantly reduces the number of new deaths (see black lines in Figure 3) and attitudes of DDC/DCC (red lines). We can see also in Table 2 that *F* / *F*_*C*_ (*k*_1_, *k*_2_) > 1 for both datasets. Large values of *F* / *F*_*C*_ (*k*_1_, *k*_2_) calculated for the relationship DDC/DCC versus VC show that vaccinated infected persons will most likely not die, i.e. the course of the disease will not be very severe.

We can use eq. (3) in order to estimate the critical values of the vaccination level VC when the new deaths cease to appear. Taking the values of *a=*4.1071 and *b=*-0.0521 corresponding to the relationship DDC versus VC (see Table 2) we obtain the critical vaccination level 78.8% for the European dataset. For the complete dataset this figure is 90.7%. The black lines in Fig. 3 illustrate these critical values. The most reliable relationships DDC/DCC versus VC yield the critical values 83.1% and 78.8% for European and complete datasets, respectively (see red lines in Fig. 3). Looking at Table 1, we see that as of September 1, 2021, no European country has reached the appropriate critical level of vaccination 83.1%. The same can be said for other countries and regions shown in Table 1.

## Discussion

It should be noted that the vaccination levels listed in Table 1 are calculated based on the full volume of populations (including children), so a further increase in VC values (in order to exceed critical figures) requires vaccination of children. Another disadvantage of existing vaccines is their ineffectiveness against new strains of coronavirus, because as mentioned above, new cases will continue to appear even with 100% vaccination. Here is a very illustrative example of Israel, which was experiencing a strong wave in September 2021, despite a fairly high level of vaccination (more than 63.3%). In particular, the daily number of new cases (DDC) exceeded the values registered before the start of vaccination, [4]. The emergence of new cases (even if they are not fatal) increases the likelihood of new more pathogenic strains, which can dramatically worsen the situation. Therefore, to overcome the pandemic, we should not rely only on vaccination.

The presented statistical analysis shows that vaccinated individuals can become re-infected and are just as dangerous to others as those who have not been vaccinated (since the DCC values do not correlate with the vaccination level). Therefore, the introduction of special passports that remove restrictions for vaccinated persons is questionable. Having fresh PCR tests can be a more effective pass to crowded places in order to prevent the spread of infection.

Many EU countries are ready to lift all coronavirus restrictions and Denmark has already done this on September 17, 2021, [11]. The obtained results allow us to estimate the risks connected with it. First, as mentioned earlier, even 100 percent vaccination does not save from the emergence of new waves. Unfortunately, the probability of death in case of infection (daily deaths per case ratio DDC/DCC) remains high due to the fact that the current levels of vaccination are less than the critical values calculated in the previous section.

For example, in Denmark, the vaccination rate VC on September 16, 2021 was 76.38% and 3 deaths caused by coronavirus were registered. Putting this figure into eq. (1) with corresponding values of coefficients *a* and *b* for European case from Table 2:

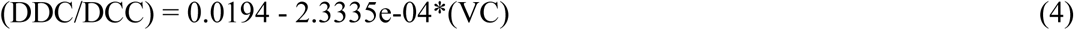

we can obtain DDC/DCC =0.0016. Thus, for every thousand new cases of infection (the number of which may become quite large), approximately 1.6 deaths could be expected in Denmark.

The results obtained in this study do not in any way deny the need for vaccinations. On the contrary, equation (4) shows that the mortality rate can be significantly reduced with a high percentage of vaccinated persons. In the example of Denmark, we see that vaccinations have made it possible to reduce mortality rate DDC/DCC by about 12 times (DDC/DCC=0.0194 at VC=0%). But, unfortunately, it will not be possible to completely stop the COVID-19 pandemic with the use of existing vaccines, since DCC values do not correlate with the vaccination level VC.

To discuss the possibilities of complete cessation of the pandemic, let us pay attention to the dependence of CC versus TC/CC for the complete data set shown by the blue dashed line in Fig. 2. As mentioned earlier, the epidemic can be taken under complete control (CC values are close to zero), if the number of tests per case is high enough (TC*1000/CC > 520).

Indeed, if every detected case is accompanied by testing of many possible contacted persons and isolation of the infected ones, then we have a chance to completely stop the spread of infection. For the case of coronavirus, the number of such tests is quite large. But as the situation in Australia shows, the appropriate level of testing can be achieved (see Fig. 2). Due to this the CC value in this country is 2193.25 which is much less than the corresponding figures for other countries and regions listed in Table 1 (for example, in the whole world, this value is 12.6 times higher).

Hong Kong and mainland China have even lower CC values: 1603.776 and 65.772, respectively (as of September 1, 2021). The test per case ratios were rather high for Hong Kong: 300.8 (January 31, 2020); 222.9 (July 31,2020); 680.4 (August 31, 2021); all figures were calculated with the use of information about accumulated numbers of tests and cases available in [1]). Thus, at the beginning of the COVID-19 pandemic, the tests per case values were less than critical one, but in the summer of 2021 they exceeded the critical level. Apparently, this allows more or less effective control of the epidemic in Hong Kong at rather low vaccination level. For example, as of September 1, 2021 DCC=0.738 which is 111 times less than worldwide (see Table 1) and VC=46%.

It seems that in mainland China, many more tests were performed per one laboratory-confirmed case. Unfortunately, the data reported by JHU [1] allows us to calculate only two values 1078 (June 24, 2020) and 1891 (August 6, 2020). Such high levels of testing are likely to have led to complete control of the epidemic in China. The value DCC=0.019 (September 1, 2021, [1]) is 4312 times lower than worldwide (see Table 1).

Increasing the number of tests requires significant costs. But in the periods between pandemic waves (when the daily number of new cases is small), even poor countries can afford to do extensive testing of possible contacts and rapid isolation of infected people. If the number of tests per case everywhere exceeds 520, we will have a chance to stop the COVID-19 pandemic.

## Conclusions

A simple statistical analysis of the daily number of new cases (DCC) and deaths (DDC) per capita showed that vaccination can significantly reduce the likelihood of deaths (DDC/DCC values). However, existing vaccines do not prevent new infections, and vaccinated individuals can spread the infection as intensely as unvaccinated ones. Therefore, it is too early to lift quarantine restrictions in Europe and most other countries.

The constant appearance of new cases due to re-infection increases the likelihood of new coronavirus strains, including very dangerous ones. As existing vaccines are not able to prevent this, it remains to increase the number of tests per registered case (TC*1000/CC values). Statistical analysis showed that if the critical value of 520 is exceeded, one can hope to stop the occurrence of new cases.

## Data Availability

data is in the text

